# Model-M: An agent-based epidemic model of a middle-sized municipality

**DOI:** 10.1101/2021.05.13.21257139

**Authors:** Luděk Berec, Tomáš Diviák, Aleš Kuběna, René Levínský, Roman Neruda, Gabriela Suchopárová, Josef Šlerka, Martin Šmíd, Jan Trnka, Vít Tuček, Petra Vidnerová, Milan Zajíček, František Zapletal

## Abstract

This report presents a technical description of our agent-based epidemic model of a particular middle-sized municipality. We have developed a realistic model with 56 thousand inhabitants and 2.7 millions of social contacts. These form a multi-layer social network that serves as a base of our epidemic simulation. The disease is modeled by our extended SEIR model with parameters fitted to real epidemics data for Czech Republic. The model is able to simulate a whole range of non-pharmaceutical interventions on individual level, such as protective measures and physical distancing, testing, contact tracing, isolation and quarantine. The effect of government-issued measures such as contact restrictions in different environments (schools, restaurants, vendors, etc.) can also be simulated.

The model is implemented in Python and is available as open source at: www.github.com/epicity-cz/model-m/releases

## 1 Introduction

Simulations using a combination of social contact network and epidemiological models are pivotal in providing solid basis for evidence-based interventions (Squazzoni et al., 2020). Epidemiological agent-based models represent an approach that simulates the epidemics on realistic synthetic population with graph describing its contacts (Eubank et al., 2004), (Hunter et al., 2018).

In this paper we present an implementation of such a model where the agents represent a synthetic population of 56 thousand people connected by a realistic network of social contacts. The network is composed of 2.8 million edges in 30 layers corresponding to various types of contacts, from families and neighbourhood to work, school and public transportation. The population and its contact network represent a detailed model of the Hodonin county in the Czech Republic. The underlying COVID-19 model is a SEIR-like model with several states representing asymptomatic and presymptomatic classes, and a set of states for detected individuals.

Unlike previous studies, we do not use stylized network structures such as random graphs (Durrett, 2010) or small-world networks (Block et al., 2020). Instead, we use sociological and epidemiological data to construct a fidel network of a mid-sized town with its surrounding area on which we simulate the spread of SARS-CoV-2.

The main utility of our multiplex approach is that each layer of the network represents a locus of interactions between individuals, such as schools for students, workplaces for individuals in productive age etc. This not only adds realism to our network model, but it also enhances the granularity of the interventions that we simulate.

The nodes are state-based agents corresponding to a SEIR-based epidemiological model undergoing simulation of epidemics spread and interventions realized by dedicated algorithms. The model allows for per-individual custom parameters based on age, sex, level of protection and other characteristics. Among the simulations of non-pharmaceutical interventions, algorithmic policies for partial closures, testing and tracing are implemented.

In the following Methods section we present technical details about creating the synthetic population and the network of contacts, the underlying epidemiological model, and the implementation of intervention procedures. An illustration of the simulation for Spring and Summer of 2020 in Czech Republic is briefly presented in the Results section.

## 2 Methods

### 2.1 Model Framework

In order for our model to be a realistic tool for estimating the effect of various interventions, we account for the following factors in the construction of the network:

1. social and geographic structure of the given town according to the latest census data
2. structural properties typical for human social networks (Rivera et al., 2010; Snijders, 2013)
3. sociological knowledge about the behavior before and during the pandemic (PaQ (2020) and MEDIAN (2020))
4. epidemiological and virological properties of SARS-CoV-2 (compartmental SEIR model (Kermack and McKendrick, 1927), (Trawicki, 2017))
5. probabilities of viral transmission of COVID-19 in each type of contact (our own expert surveying)

Our model builds upon a multiplex or multilayer network (Kivelä et al., 2014), in which individuals in a given population (such as a town) are represented as nodes, and interactions among them are represented as different layers of edges according to the environment, in which given interactions occur (e.g., family, workplace, schools etc.).

5not use stylized network structures such as random graphs (Durrett, 2010)

### 2.2 Epidemiological model

The core epidemiological model is based on a standard SEIRS model (Bailey et al., 1975). The core model itself works with a set of nodes (individuals, agents), each node being exactly in one state at each time step. The time iterates in days (one tick per day), each iteration the node can change its state.

The set of states used in our model is listed in Tab. 1. All possible state transitions can be seen at Fig. 1.

**Figure 1:**
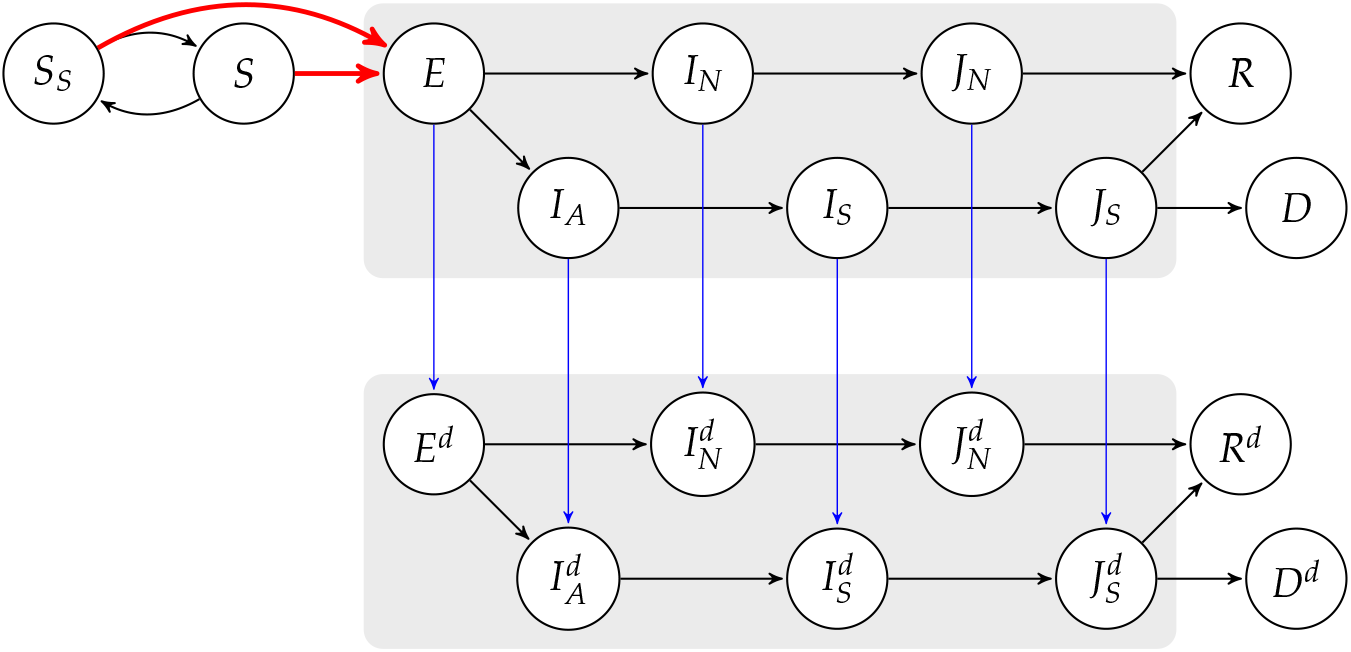
State transition diagram. See Tab. 1 for explanation of state labels. Possible transitions are depicted by arrows, where the blue arrows stand for transitions caused by being tested, the red ones for being exposed to virus (having been infected).

There are three types of transitions. First, those depicted by black arrows, happen on the daily bases with fixed probability (see Section 2.3 for the matrix of transition probabilities). To set up these probabilities, we assume the occupation time in a state to be exponential and set the day-by-day transition probabilities accordingly (so the model approximates a continuous-time Markov chain by a discrete Markov process), see Tab. 2 for parameters used for deriving corresponding density distributions.

For example, the occupation time of *E* is assumed to be exponential with mean *m*_*E*_ = 5.08. Upon the end of the *E* period, the individual becomes asymptomatic (*I*_*n*_, with probability *p*_*n*_ = 0.179) or presymptomatic (*I*_*a*_, with the complementary probability). In the former case, the individual remains infectious asymptomatic for the exponentially distributed time with mean 8, in the latter case he is presymptomatic for the exponential time with mean 4, etc.

Second, blue arrows, represent transitions forced by testing. Each day, a node may be tested with probability *θ*. For the simplicity, we keep *θ* non-zero only for nodes exhibiting symptoms.

If node is tested, it changes its state to its detected counterpart. To better control the portion of nodes being tested, we also keep the test rate parameter *τ*. As soon as a node starts to exhibit symptoms, it sets its *θ* to non-zero number with probability equal to *τ*. A node can be forced to move to detected state also via quarantine and contact tracing policy module (described in Section 2.6).

Last, the red arrows represent state transitions induced by being exposed to the virus (with the consequent illness). The calculation of probability that this transition happens is based on a multi-graph *G* that is a set of edges between the nodes of the model. Each edge represents possible contact in the region represented by a graph. Between every two nodes, there can be zero, one or more edges. An edge *e* is identified by its layer type, sub-layer type, probability *p*_*e*_ of contact and intensity *p*_*e*_.

Every iteration (day), for every edge *e*, an imbalanced coin is flipped with probability *w*_*l*_ *p*_*e*_, where *w*_*l*_ is a weight of the *e*’s layer *l*. Then, according to the result of the toss, *c*_*e*_ is set to 1 (there is a contact on the edge), otherwise *c*_*e*_ = 0.

The (conditional) probability of transition from *S* or *S*_*S*_ to *E* for a node *n* (that is in one of *S*-states) is then given by

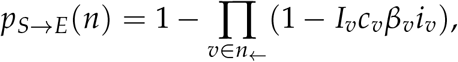

where {*v*; *v ∈ n*_*←*_} is a set of edges adjacent to the node *n* and *I*_*v*_ is one if node *x*, where *v* = (*n, x*), is infectious (i.e. in one of states *I*_*n*_, *I*_*a*_, *I*_*s*_), zero otherwise; *β*_*v*_ is a model parameter of infectiousness (and may depend on the layer type and whether the infectious node is symptomatic or not). See Appendix 2.4 for the details on how the graph is constructed and Appendix 2.5 for the way weights of the layers were determined. Further, see Section 2.3 for the transition probabilities between the states except for the transitions to *E*, which has been described above.

The described computation core is encapsulated in a framework performing simulation of state policies. These are quarantine and isolation, contact tracing, testing, limiting contacts and restricting access to public places, such as closing schools. Site closures are implemented by changing weights of corresponding layers. Quarantine and isolation are realised by adjusting edges’ parameters for individual nodes isolated nodes.

In addition, the model follows a calendar of parameters and accordingly changes layer weights (according active site closures) or various model parameters (such as reducing *β* if masks are used, adjusting test rate and *θ* according testing capabilities). This calendar for the Czech Republic is provided by (PaQ, 2020) and (MEDIAN, 2020) and follows the real situation in CR. See Section 2.6.1 for the way of computation the contacts reductions.

Afther each iteration (on daily basis) the core model activates the policy module that controls the mentined parameter changes and simulates quarantine, isolation and contact tracing policies by modifying the graph.

The more details on policy modules, quarantine and contact tracing simulations can be found in Section 2.6.

### 2.3 Epidemiological Parameters and Transition Matrix

In the present Section, we list parameters of our Epidemiological model. In Table 2 some parameters of the COVID illness, surveyed from the literature can be found.

**Table 1:** List of possible states. Upper index *d* stands for detected variant of the state (a node was detected). The typical node life cycle is *S → E → I*_*n*_ *→ J*_*n*_ *→ R* for individual with asymptomatic progression and *S → E → I*_*a*_ *→ I*_*s*_ *→ J*_*s*_ *→ R*/*D* for individuals exhibiting symptoms during illness.

|  |  |
| --- | --- |
| $S$ | susceptible |
| $S_s$ | false symptomatic (with symptoms but not infected by COVID) |
| $E, E^d$ | exposed |
| $I_a, I_a^d$ | infectious presymptomatic (later will exhibit symptoms) |
| $I_s, I_s^d$ | infectious symptomatic |
| $J_s, J_s^d$ | non-infectious symptomatic |
| $I_n, I_n^d$ | infectious asymptomatic (never will become symptomatic) |
| $J_n, J_n^d$ | non-infectious asymptomatic |
| $R, R^d$ | recovered |
| $D, D^d$ | dead |

**Table 2:** Epidemiological parameters

| Par. | description | value | error | Source |
| --- | --- | --- | --- | --- |
| $m_E$ | expected occupation of $E$ (incubation period) | 5.08 | 95%CI 4.77-5.39, SD 0.18 | He et al. (2020) |
| $m_i$ | expected duration of infectiousness | 8 | | Wölfel et al. (2020) |
| $m_a$ | expected duration of presymptomatic period | 4 | IQR2-7 | Nie et al. (2020) |
| $m_s$ | expected duration of RNA positivity for symptomatic | 25.2 | SD 4.9 | Noh et al. (2020) |
| $m_n$ | expected duration of RNA positivity for asymptomatic | 22.6 | SD 4.0 | Noh et al. (2020) |
| $p_n$ | probability of completely asymptomatic progression | 17.9 | 95%CI 15.5-20.2 | Mizumoto et al. (2020) |
| $c$ | case fatality rate | 0.018 | 95%CI 0.0118-0.0243, SD 0.0032 | He et al. (2020) |
| $f$ | false symptoms rate | 0.0003 | | NIPH (2020) |
| $m_f$ | false symptoms duration | 5.5 | | NIPH (2020) |

Next we give the matrix of transitions between individual states. For space reasons and clarity, we split the matrix into that between “undetected” states (Table 3) and that between “detected” ones (Table 4). Note that the matrix includes only “natural” transitions, not those caused by detection, quarantine etc. On the diagonal of the matrix, there is always a number such that the row of the matrix sums to one.

**Table 3:**
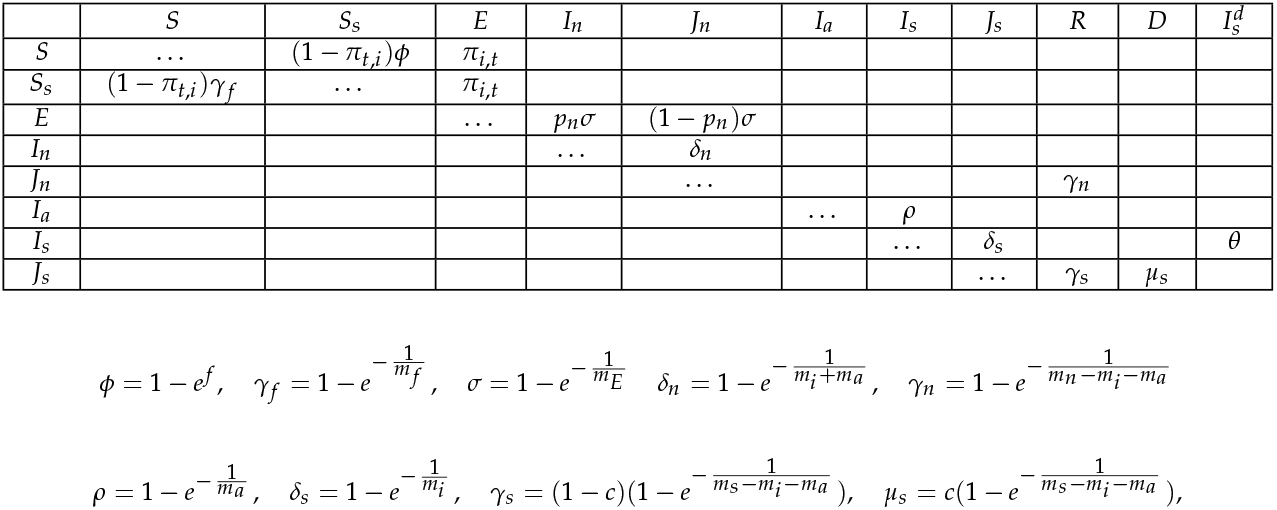
Transitions between “undetected” states

**Table 4:** Transitions between “undetected” states

| | $E^d$ | $I_n^d$ | $J_n^d$ | $I_a^d$ | $I_s^d$ | $J_s^d$ | $R^d$ | $D^d$ |
| --- | --- | --- | --- | --- | --- | --- | --- | --- |
| $E^d$ | ... | $p_n\sigma$ | $(1 - p_n)\sigma$ | | | | | |
| $I_n^d$ | | ... | $\delta_n$ | | | | | |
| $J_n^d$ | | | ... | | | | $\gamma_n$ | |
| $I_a^d$ | | | | ... | $\rho$ | | | |
| $I_s^d$ | | | | | ... | $\delta_s$ | | |
| $J_s^d$ | | | | | | ... | $\gamma_s$ | $\mu_s$ |

### 2.4 Construction of the Contact Graph

We consider a region consisting of a middle-sized town Hodonín (about 24 thousands of inhabitants) together with a selected surrounding municipalities (additional 32 thousands). We had three basic data sources describing the region at our disposal. First, it is the list of inhabitants of the region according to the 2011 census, each record including the age, gender and the municipality they live in, the records being grouped into so called economic households (those having common housekeeping). Second we have the list of houses including their type (family or apartment building), their location and the number of apartments included. Third, we have the municipality level data coming from the 2011 census about the economic activity class (students, workers, housepersons), industry sector if working, and the information whether and how long the person is commuting. The additional data sources, used for the graph construction, include public surveys PaQ (2020), (MEDIAN, 2020), (MEDIAN, 2019), a study CDV (2020), Points of Interest list by the Eoconlab company, and expert estimates of shop cappacities by Zdeněk Skála.

#### 2.4.1 Population reconstruction

As the mentioned three data sources – the census person level data, the house data and the additional municipality-level aggregate census data – are not mutually connected, we randomly assigned the individuals their economic activity and commuting status (both with respect to their gender and age), and randomly assigned their households into the apartments.

In particular, when “inhabiting” the apartments, we first randomly “occupied” all the apartments (with the exception of two small villages where there were less households than apartments, so we left some apartments free), and, consequently, we randomly assigned the remaining households into the available family houses. While there were little of these excess households in the town, there were 1.5 to 2 times more households than apartments in the smaller municipalities, so two households were assigned to a majority of the apartments there. To replicate the fact that distinct generations often live in family houses, we refused with probability 0.9 assigning an additional household into an apartment whenever the difference of the average age of the households was less then 20.

Next, we randomly assigned the economic activity to the individuals as follows: for each individual, we performed a Bernoulli trial with the parameter equal to the probability that a person of a given age and gender is working, computed from the aggregate census data. When the result was “working”, we randomly assigned the person an industry sectors he/she is working at according to the distribution given by the aggregate data; these sectors are listed in Table 5. If the result of the trial was “not working” and the individual was in school age 6–18 (both included), then we made him/her a student; if his/her age was 19–24, then we tossed a coin with probability 0.6 for woman, 0.7 for man, whose heads made the person a student. In all the other cases, we classified him/her as house-person, if she was under 65, or retired otherwise. The age constants and the probabilities were chosen in order the resulting distribution to replicate the municipality data.

**Table 5:**
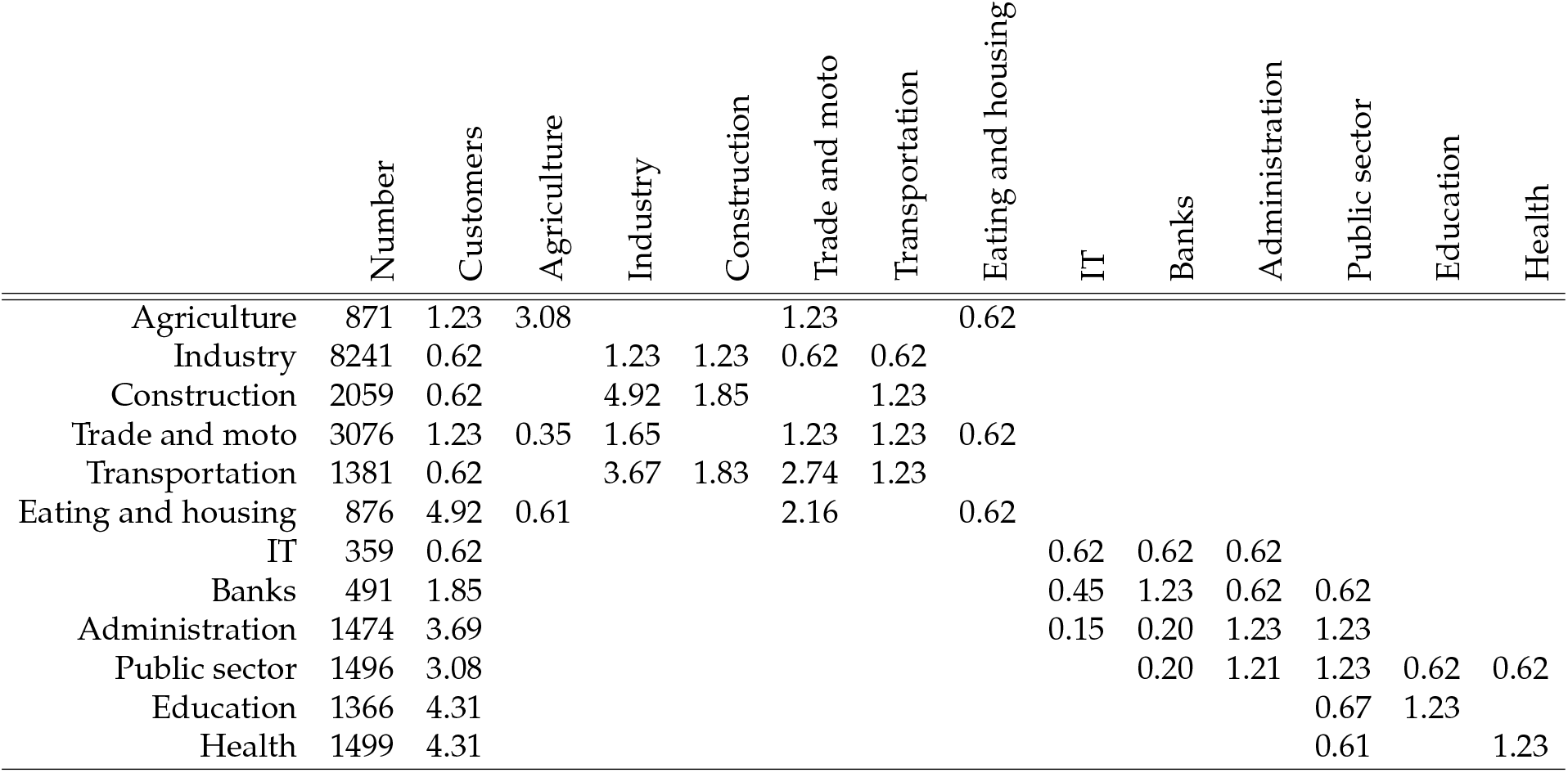
Work contact intensities

Next, according to the distribution of the aggregate data, which is available separately for woman, men and students, we randomly assigned each person one of the commuting levels: not commuting (i.e. not leaving the municipality for work or study), commuting less than 15, less than 30, 45, 60, 90 or more minutes.

As a result, we roughly replicated all gender, age, economic activity and daily mobility of the region’s population, as well as its geographical structure.

#### 2.4.2 Social contacts

In a usual matter, we modelled the social network of the population by an undirected graph, with the individuals as nodes and the potential contacts as edges. In line with (Mossong et al., 2008), as a contact we regard either a two-way conversation with three or more words or a skin-by-skin contact (e.g. handshake). We assumed that, during a single day, any potential contact happens with a certain probability *p*, and, given that the contact happens, the contagion takes place with another probability *ι*.

It is needless to say that estimating human contacts is an extremely complex task dependent on a vast number of unknowns. One way of approaching this problem could be to assume the contact model simple enough so that it could be calibrated by available scientific evidence, the other way could be to do it arbitrarily, according to “common sense”. Yet the first option seems safe, precluding “self-fulfilling prophecies” (i.e. “what we assume that we get”), we did not go purely this way because, if we did so, the added value of modelling the network person-by-person would be lost.

We give an example: As there is no study on detailed structure of work contacts to our best knowledge, it would be “scientifically fair” to assume that these contacts form a random graph with degrees, corresponding to the (known) age structure of the contacts. However, in reality, workers usually meet in small groups (e.g. offices), which fact cannot be reproduced by a random graph. On the other hand, a construction of a graph involving these groups naturally brings questions about its parameters, the sizes of these groups in the first place; not having any scientific evidence in this respect, we are forced to set these numbers arbitrarily. As our desire is to give quality scientific predictions, but simultaneously we realize that the added value of random graph models in comparison to the compartment ones is not high, we try to keep reasonable balance between the “scientific” and the “arbitrary” approach when constructing our contact network.

Another challenge in this respect is the construction of a model of random encounters in public places such as pubs, shops or school classes. It is clear that the expected number of contacts of an individual will grow with the number *n* of the other people present; however, hardly it can grow linearly. To simplify things, in each of these situations, we assume that the number *N* of the contacts an individual is intended to realize is Poisson distributed with mean *λ*, which we call contact rate, while the number of actual contacts is min(*N, n*), i.e. a random variable with a censored Poisson distribution. As we do not prefer any contact before another, necessarily the probability of contacting a pre-chosen individual comes out as 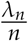 where *λ*_*n*_ = 𝔼 min(*N, n*).^*^

For instance, when going alone to a crowded pub, we can guess that a person makes *λ* = 3 contacts (say that the tables are four-seat). However, if there is only one extra guest present, then the maximum number of contacts is clearly one; thus, according to our assumptions, the mean number of contacts in that case is 1 *×* P[*N ≥* 1] = (1 *− e*^*−*3^) = 0.95.

A clear advantage of this simple approach is that only a single parameter - the contact rate - has to be set for each of these situations. However, in some cases like large supermarkets, our model has to be refined – yet it is reasonable to think that I experience, say, 3 close contacts in a crowded supermarket (two in a line and one perhaps when asking where frozen chicken are), it is unlikely, that two people meet when they are the only ones present in the supermarket; thus, in these situations, the censoring term has to be scaled by some *α <* 1 so that the actual number of contacts would be min(*N, αn*). Finally, if the other individuals attend the place only with probabilities *p*_1_, …, *p*_*n*_, then we, mildly violating the probability calculus, assume the actual number of the contacts to be 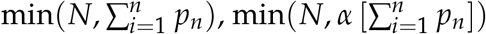 respectively.^†^

If more is known about the situation when people meet, then we proceed more explicitly. In restaurants, for instance, it is clear that the guest will have a word contact with a service, perhaps less intense than sitting at the same table, and similarly in shops. In both these cases, we assume that there is a single contact of each client with a randomly chosen member of the staff.

Another topic little supported by the available data is the geographical preference, namely the question what role a geographical distance has in choosing shops, restaurants, friends, etc. In these situations, we assume a simple random choice model in which the probability of choosing particular object is proportional to *w*_*ρ*_(*d*) = (1 *− ρ*)^*d*^ where *ρ* is the spatial preference parameter and *d* is the distance from the decision maker to the object in kilometres.

For the construction of the friendship contacts network, we wanted to respect the known properties of human friendship networks. These properties are average degree, degree distribution, assortativity (on age and gender), and clustering. The average degree (i.e., the average amount of friend contacts) is N, and its distribution is positively skewed, which means that there are some highly sociable individuals in this layer. This is generally in line with previous research (AddHealth, Rivera et al., 2010; Snijders, 2013). For friendship networks, it is also typical that these networks display assortativity on gender and age - nodes are much more likely to have ties to those of the same gender or of similar age (Rivera et al., 2010; Snijders, 2013). Lastly, a common feature of human social networks that we wanted to reproduce in this layer is clustering. Clustering refers to the tendency of connected individuals to share common friends, represented by triangles in the network (Newman, 2003, Rivera et al., 2010). To meet these requirements, we generate this layer of the network by a modified Barabasi-Albert procedure; the modified version of the procedure is used in order its results to replicate the required assortativity and clustering, and to avoid nodes having unreasonably high degrees (comparable with the population size).

In particular, for each *i ∈* ℕ, having constructed a (temporary) graph for *i −* 1 nodes, we assign the *i*-the a random out-degree *D*_*i*_ *∼* Po(6). Consequently, we generate 2*D*_*i*_ potential connections to {1, …, *i −* 1} by the Barabasi-Albert scheme (i.e. with probabilities of choice proportional to their in-degree plus one). Then, we randomly choose *D*_*i*_ edges out of the 2*D*_*i*_ potential ones with the probability of choice of the *j*-th one proportional to

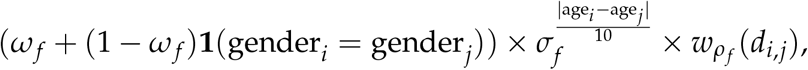

where *d*_*i,j*_ is a distance (of the apartments) of *i* and *j* in kilometres and *ω*_*f*_ = 0.2, *σ*_*f*_ = 0.3 and *ρ* _*f*_ = 0.65 are parameters, manually chosen so as to produce the required assorativity. Finally, we add connections to the selected nodes and, to enforce clustering, we randomly generate one connection between the nodes we newly connected with *i*. To avoid nodes with their degree measured in thousands, which are usually generated by the plain Barabasi-Albert procedure, we tossed a coin with 0.99 probability before adding each edge into the final graph; once tails fell, we stopped adding edges to the target node (with the B-A. scheme working as if this “censoring” did not take place).

To reflect the fact that the number of actual contacts clearly depends on the number of potential ones sub-linearly (although I can have one hundred friends, I can hardly see them all during one day), we made an assumption that the probability of a potential contact (*i, j*) happening is

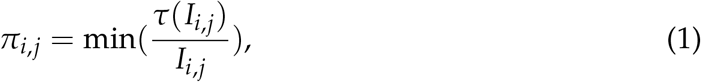

where *τ*(*x*) = *κ*(1 *−* exp{*−κx*}) for some *κ* and *I*_*i,j*_ = max(*δ*_*i*_, *δ*_*j*_), where *δ*_*i*_ is the degree of *i* (i.e. the number of its potential contacts). Again, there is only a single parameter *κ* here, which we set to *κ* = 4 in order to have the expected number of leisure contacts equal to *d*_*L*_ ≐ 2; this number was chosen because, by (Mossong et al., 2008), Figure 2, leisure contacts form about 15% of total contacts, which are 13.52 contacts by overall contact matrix. However, it should be noted here that, besides *d*_*L*_, the epidemiological predictions based on the model may be sensitive to the choice of function *τ* determining the dependence of the actual contact numbers on the potential ones, consequently affecting the infectiousness of the nodes with very large numbers connections (superspreaders).

**Figure 2:**
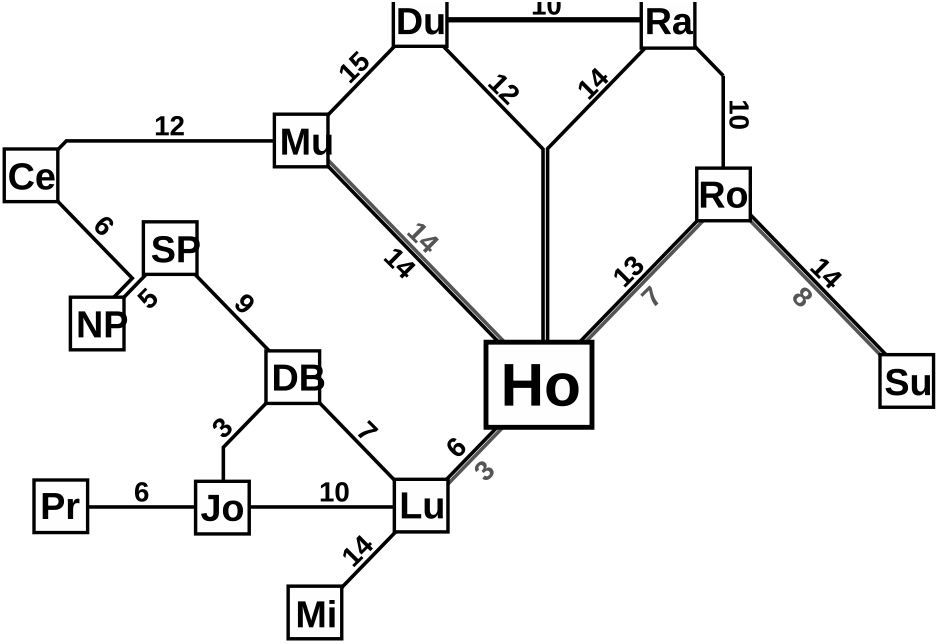
Schematic map of the region with transport connections. Numbers denote travel time in minutes. Red lines – train, black lines – bus.

In our contact network, we consider several layers of contacts, labelled *family, school, work, pubs, visits, outdoor, shopping, services* and *other*. These categories further divide into layers listed in Table 9. For each layer, we construct a separate (undirected) graph, with nodes corresponding to individuals and edges corresponding to contacts, each edge being “weighted” by a its probability *p*_*i,j,k*_ where *i, j* are connected nodes and *k* is the layer index. While edges cannot be duplicated within the layers, two nodes may be connected in different layers (e.g. work together and then go for a drink). Next we discuss the individual categories.

#### 2.4.3 Family

Having detailed (anonymized) data about the structure of the households at our disposal, we might model family contacts explicitly. In particular, we assume that each member of a household potentially meets all the remaining members; to replicate the prevalence of same-age contacts and the larger intensity of home contacts by seniors implied by the “home” contact matrix from (Prem et al., 2017), we assume that the probability of each such meeting is one if either the age difference is less or equal to ten, or one of the persons to contact is 60 years old or more; otherwise, the contact probability is set to *p*_*h*_ = 0.745. Further, we assume that, once two households live in the same apartment, any member of the first one and any member of the second one have a contact with with probability *p*_*y*_ = 0.397. Finally, we assume that each person aged 50 or more has contacts with households of their children, number of which we draw from a discretized censored normal distribution with mean 1.92 and standard deviation 0.7. i.e. the values from European Social Survey round 9 (NSD Norwegian Centre for Research Data, 2018). If two (or more) persons over 50 live in the household, we assume them to have contacts with the same “young” households. The childrens’ household we drew randomly from those, in which at least one member is aged less than the average age of the (grand)parents age minus twenty.^‡^ We assume that each member of an “old” household visits a given “young” one with probability *p*_*o*_ = 0.297 divided by the number of his children, and each member of a “young” household visits the “old” one with probability *p*_*y*_ = 0.373. During each visit, contacts of the visitor with each member of the visited household were assumed. The values of *p*_*h*_, *p*_*a*_, *p*_*o*_, and *p*_*y*_ were determined by a grid search so as to minimize the weighted square distance^§^ of the resulting contact matrix and the home matrix from (Prem et al., 2017), assuming that, in addition to the family contacts, friends visits result in home contacts (see 2.4.6).

#### 2.4.4 School

As we had the list of schools, including their location and capacity, at our disposal, we could model the school network explicitly. We proceeded as follows: according to their age, we divided the children-aged individuals into those attending kindergartens (those with age 3 to 5), lower elementary schools (6 to 10), higher elementary ones (11 to 14) and highschools (15 to 18). As there is no university in the small town we deal with, we assumed all the older students travel outside for study. Next we randomly reordered all the students’ lists and, for all but the highschool list, we gradually assigned each student to the nearest school which has not yet reached its capacity. As in reality, not all kindergarten candidates could be placed due to capacity reasons while all the elementary candidates did. For the highschool candidates, the distance did not play role; instead, the students were assigned their schools randomly. Similarly to the elementary schools, all highschool candidates could be placed not overrunning the declared capacities of the schools.

Further, in each school, we distributed the students into classes. Assuming that students of the same age form the same grade, we uniformly split each grade into classes no greater than 20 students. Next, for each class, we found a class teacher among the individuals working in *education* sector the following way: First we (randomly) searched among those living in the same municipality and, after we eventually run out of them, we chose a random one from those who commute less than 30 minutes, which is the time the majority of travels within the region can be made in. Having found the class teachers, we “hired” twice as much additional teachers for the school the same way.

The contacts of students within classes, outside the classes and the mutual contacts of teachers were modelled as it was described at the start of the present Section, assuming contact rates *λ*_*s*_ = 4.72, *λ*_*o*_ = 1.88 and *λ*_*t*_ = 0.777, respectively. The contact rate of a class teacher and his class was set to *λ*_*cs*_ = 1.44 meaning that the actual number of the contacts of that kind will be min(*N, n*), *N ∼* Po(*λ*_*cs*_) where *n* is the number of students in the class. The contact rate of any teacher and any student of the school was set to *λ*_*ts*_ = 0.444. All the five contact rates were determined so as to match the school contact matrix by (Prem et al., 2017) similarly as with home contacts.

#### 2.4.5 Work

As it was described in Subsections 2.4.4, and will be described in Subsections 2.4.6 and 2.4.7), work contacts of teachers, restaurant staff, salepersons, respectively, were constructed explicitly. However, these contacts form only about 4 per cent of work contacts, given by the “work” contact matrix from (Prem et al., 2017). In the present Subsection, we describe the way we construct the remaining majority of contacts.

Lacking any relevant study identifying a distribution of work contacts into those takes place in the workplace between workers, those between sectors and those with customers/clients, we estimated the frequency and structure of these contacts based on the survey data provided by PAQ (PaQ, 2020), see Table 5; the matrix is normalized so that the average number of working contacts equals 6.04, which is the average number of work contacts given by the “work” contact matrix from (Prem et al., 2017). Note that the matrix is asymmetric, reflecting different numbers of workers in different sectors.

In creating the working contact network, we proceeded as follows. First, we randomly assigned the workers commuting within the region (i.e. those commuting up to 30 mins) to individual municipalities with the choice probabilities proportional to the municipalities’ population. Further, having all workers assigned to municipalities, we, slightly abusing reality in which workers travel during their working hours, created a separate working network for each municipality.

Each network was created the following way: First, for each sector, we randomly determined the number and the sizes of workplaces, drawing each from the Poisson distribution with mean equal to twice the expected number of contact rate within the sector (see the diagonal of Table 5). For each sector, we created as much workspaces as it was needed to cover the number of the workers in the sector (the size of the last one was possibly truncated). After randomly assigning the workers into the workplaces, we created the network of mutual one-probability contacts within each workplace with the contact rate equal to the minimum of the corresponding diagonal value and the workspace size.

Next, for each worker and each sector except for her own, we created *R* random one-probability contacts where *R* is Poisson with mean equal to the half of the expected inter-sector contact rate (the off-diagonal values in Table 5) multiplied by an age-dependent weight proportional to the expected contact rate of the corresponding age category from the “work” contact matrix from (Prem et al., 2017). In choosing counterparts of these contacts, the choice probabilities were age-weighted, too.

Finally, for each worker, we drew the number of his contacts to the customers from Poisson distribution with intensity 10*×* the corresponding value from the second column of Table 5, and assigned each contact 0.1 probability of happening. The counterparts of the contacts were chosen randomly from all the population of the region. For simplicity, we assumed that these contacts always happen in the same municipality as the workplace lies.

#### 2.4.6 Leisure

In our construction, we assume three types of leisure contacts: visits at home, going to restaurant and an outdoor activity.

In particular, for each two nodes *i, j* connected in the friend network such that both *i* or *j* are at least 18 years old, we draw from a Bernoulli variable with an age– and gender-dependent probability of going to a restaurant in the evening, computed from (MEDIAN, 2019). Given the positive result of the trial, we randomly found a restaurant near to the apartment of *j*; in particular, the restaurant was chosen from the distribution with probabilities proportional to 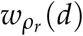 where *ρ*_*r*_ = 0.5 is the spatial preference and *d* is the distance of the restaurant from the apartment of *j*.

In the case the “restaurant” coin fell tails or any of the parties in the contact was younger than 18, we tossed another coin with *p*_*v*_ = 0.5, heads saying that *i* visits *j* in the *j*-ths home; tails meaning that *i* and *j* are going to carry out some outdoor activity. Not having any relevant data in this respect, we determined *p*_*v*_ by guess.

In all the three cases (restaurant, visit or outdoor), a contact of *i* and *j* with probability *π*_*i,j*_ was added to the corresponding layer (see (1)). In the case of home visit, the contacts of *i* with all the members of the *j*-th household were added (with the same probability). Finally, having constructed all the restaurant contacts, mutual contacts of each restaurants guests with contact rate *λ*_*u*_ = 2 and contacts of guests and two-member staff with rate *λ*_*pg*_ = 1 were added. Both *λ*_*u*_ and *λ*_*pg*_ were determined by guess, the first reflecting the fact that four-seat tables are common in restaurants, second one stressing the payment as the only longer contact of a guest with a waiter.

#### 2.4.7 Shopping

Having estimates of shopping behaviour from (MEDIAN, 2019), the list of the shops in the region and expert estimation of the turnout for the largest shops, we could model shopping explicitly to some extent.

We proceeded as follows: First, as the list of shops we use as input does not distinguish their size, we classified the shops visually (using Street-view of Google maps) into three categories: small shops, self-service shops and super/hyper-markets. Then, for each person aged 18+, we tossed three coins with the age-category, gender, and economic activity dependent probability of visiting each category of shops given by (MEDIAN, 2019). Given each heads, a shop of the corresponding category was randomly chosen with spatial preference 0.6, 0.4, 0.2 respectively. For the five biggest super– /hypermarkets, for which we had expert turnout estimates at our disposal, their choice probabilities were manually adjusted to reach comparable turnouts in the simulation.

For each shop, we assumed contact rates of its customers to be *λ*_*c*_ and scale 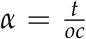 where *t* is the average shopping time, *o* is the opening time and *c* is the number of cashiers. We assumed staff number of each shop to be *s* and a staff–customer contact rate to be *λ*_*f*_. The values for individual types are listed in the Table 6.

**Table 6:**
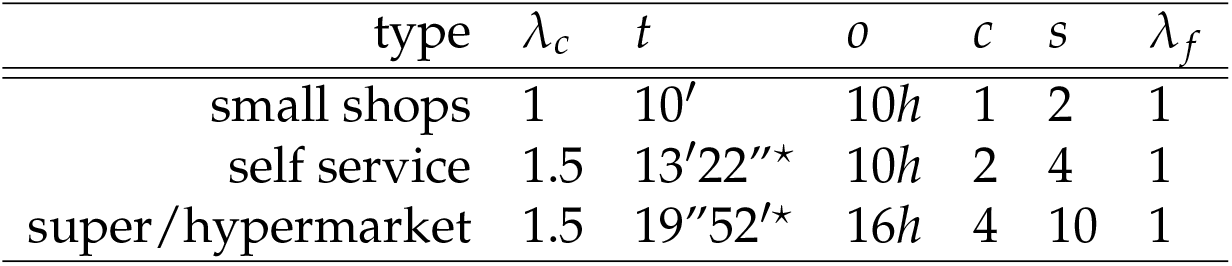
Shop parameters. Starred values denote external expert estimates, the rest being our guesses.

#### 2.4.8 Other contacts

Finally, we generate contacts purpose of which is unknown to us. For each individual from the population, we generated three potential contacts of this kind, each with probability of happening *p*_*o*_ = 0.27. Following the uncertainty principle, we generated these contacts randomly over all the network; however, to replicate the obvious diagonal of the “other” contacts matrix from (Beaumont, 2010)prem2017projecting, we weighted the choice probabilities of an *a*-aged node by 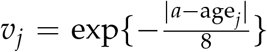. Whenever the parties of a contact were found to live in different municipalities, we tossed a fair coin to determine, which party will travel to the other one (see Subsection 2.4.9). The value of *p*_*o*_ was set manually so that the average number of “other” contacts matches with that determined by the “other” matrix from (Prem et al., 2017).

#### 2.4.9 Transport

As it is clear from the previous text, in our model, individuals travel for various reasons: commuting (i.e. regularly going to school and work), family visits, friend contacts, shopping travels, customers’/clients’ travels for services, and travels associated with contacts from the category *other*.

The travelling is modelled as follows. Whenever an individual is commuting inside the region for work or school, a two-way trip from his home to his workplace is generated. Further, a two way trip is generated whenever a family– or friend visit takes place with the visitor living in a different municipality than the visited one. Similarly, a two-way trip is generated on occasion of shopping or use of services outside the home municipality. Finally, a two-way trip is generated for each contact from the *other* category outside a municipality.

In determining the public transport contacts, we proceeded as follows. For each generated (two way) trip we tossed an asymmetric coin^¶^ determining whether the trip will be done by means of public transport. Next, for each pair of destinations, we determined the list of sections which will travellers go through by the shortest-path algorithm. Thus, we could determine which individuals will potentially travel through each section. Finally, we generated contacts (possibly) happening in each section, assuming contact rate *λ*_*u*_ = 0.5 and scaling *α*_*u*_ = 0.25. The values of *λ*_*u*_ was determined so as to roughly replicate the ratio of transportation contacts seen in (Mossong et al., 2008), Figure 2. The scaling factor was roughly given by an average number of connections per day in the involved lines.

#### 2.4.10 Validation

As it was already mentioned, our model has very many “degrees of freedom”, so it is very difficult to construct it purely scientifically, using only “objective data”. In the present Subsection, we discuss how “objectively funded” the individual parts of our model are.

In construction of our model, our main “objective anchors” are the contact matrices from (Prem et al., 2017). The fit of our contact matrices with them is reported in Table 7 and graphically illustrated in Figure 3.

**Table 7:** Fit of the contact matrices to (Prem et al., 2017)

|  | Matrix | home | work | school | other |
| --- | --- | --- | --- | --- | --- |
| Expected contacts |  | 3.39 | 6.03 | 6.02 | 4.42 |
| (Prem et al., 2017) |  | 3.36 | 6.07 | 6.02 | 4.45 |

**Figure 3:**
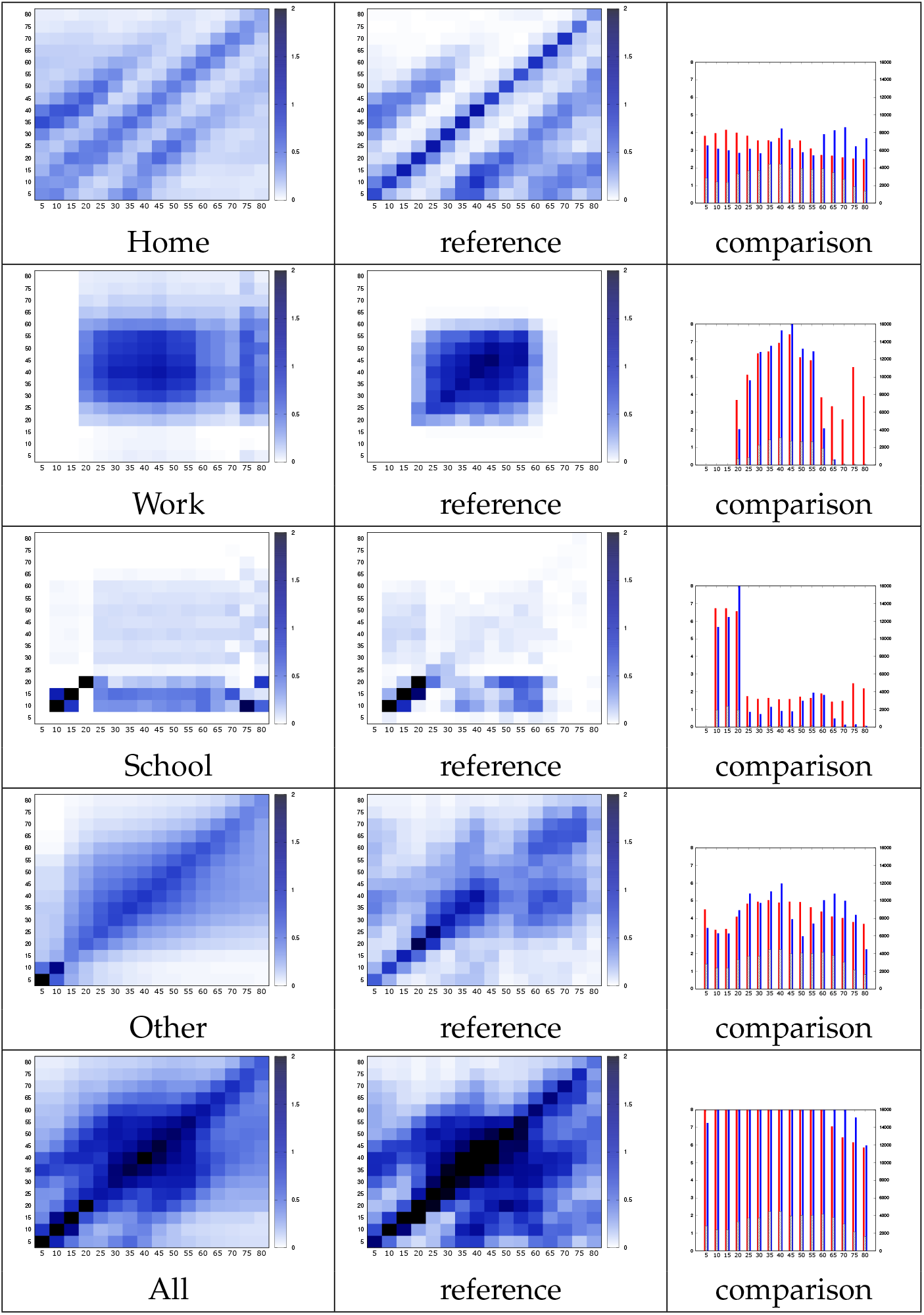
Fit to (Prem et al., 2017). Right column: blue: our value, red: reference value, hatched: population (weight)

As we could directly compare age-specific contacts from *family, work*, and *school* category with corresponding reference matrices, we could be “quite sure” with these categories. Unfortunately, this is not true for the remaining categories because their are all aggregated in the “other” reference matrix. so we can validate their parameters only indirectly.

First of all, we cannot be sure of the proportion of contact categories within the friend network. Yet we have probabilities of visiting a restaurant, we do not know how many takes place there. Similarly, we cannot know the proportion of household visits and outdoor (and perhaps other) activities, not even thinking about the clear seasonal change of this proportion. As the leisure contact categories: *pubs, visits*, and *outdoor*, form a significant portion of the contacts (see Figure 4), we should be slightly cautious when e.g. evaluating the impact of restaurants visits to the virus spread.

**Figure 4:**
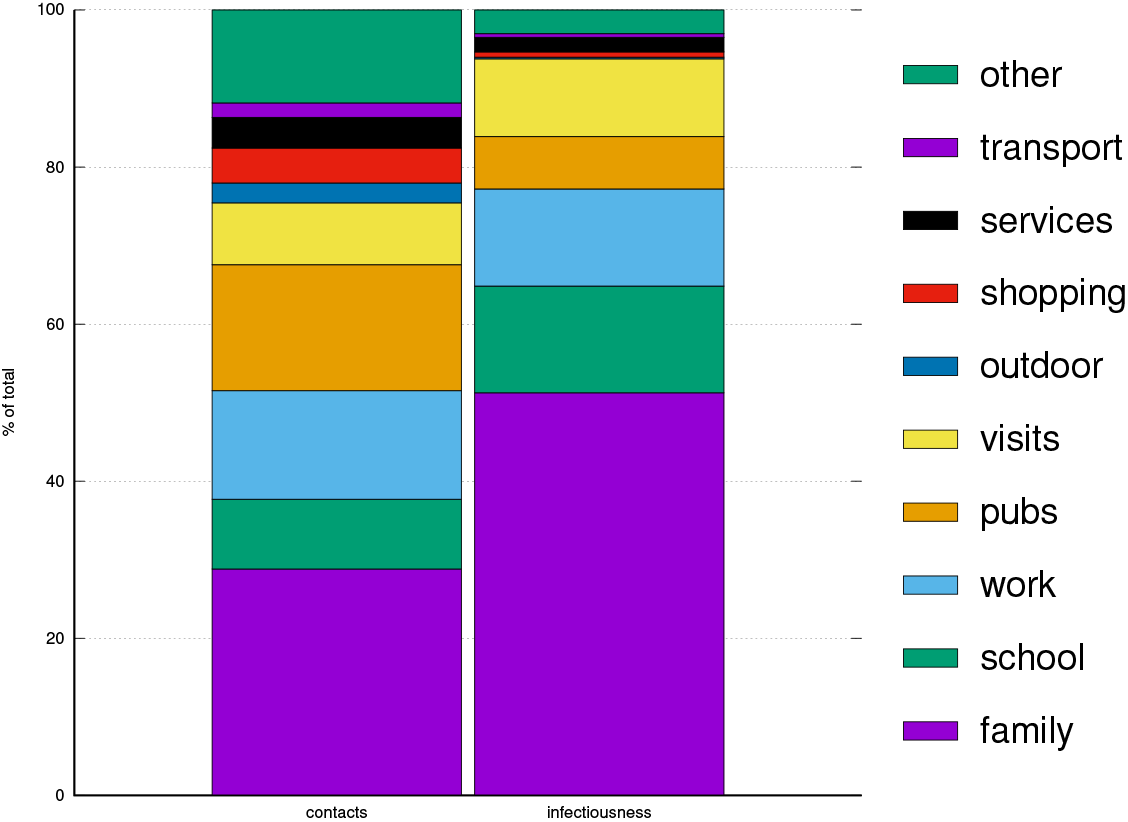
Ratio of expected contacts and infectiousness by category

The situation is similar with shopping: yet we can estimate the number of shop visits thank to the shop visit probabilities from (MEDIAN, 2019), we can only speculate how many significant contacts people have while shopping; however, as these contacts are usually short, the significance of possible misspecification here would hot be severe as in the case of leisure contacts.

Finally, there is a great uncertainty regarding the nature of contacts grouped in the *other* category, further introducing the uncertainty into the the relevance of this category with respect to infectiousness (see Subsection 2.5 as well as the right column of Figure 3).

Moreover, there are parameters of our model, which are not reflected in the contact matrices. The most important of them are geographical preferences. Fortunately, these parameters can be verified at least indirectly via the public transportation. Yet it plays little role in our simulation (see Figure 4), which is mainly due to the size of the town and small duration of contacts in transport, we can check whether our numbers of travellers correspond to the capacity of the public transport network. The results of this check can be found in Table 10 in which the expected numbers of travellers stemming from our simulation are compared with the public transport capacity in individual travel sections.^∥^ Apart from two less populated sections, the results seems realistic. Yet one can bear in mind that travelling outside the region is not included here, co the actual numbers would be higher, it can be concluded that our spatial preferences assumptions are not completely unrealistic.

The summary results of our simulation may be seen in Figure 4 and Table 9.

### 2.5 Contagion Probabilities

For a contact of *i* and *j* on layer *k*, we assume that the probability of *j* infecting *i* at time *t* through this contact provided that the former is infectious (see below) is

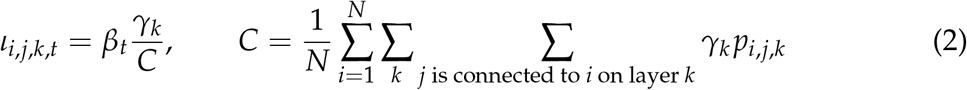

where *β*_*t*_ is a constant, dependent only on time, *p*_*i,j,k*_ is the probability of contact of *i* and *j* on layer *k* and *γ*_*k*_ is a layer-specific constant. The normalization by *C* is done in order *β*_*t*_ to be comparable with its analogy in compartmental models, in which, hypothetically, a single contact happens each period. Due to the normalization, clearly, it suffices for *γ*_*•*_ to be determined only up to a common multiplicative constant (i.e. only ratios of *γ*_*•*_ matter).

We consider eight possible levels of the contacts infectiousness (hence eight possible values of *γ*_*k*_), see Table 8. Due to the lack of knowledge about the infectiousness of the COVID virus, we chose a non-standard way of determining the value of *γ*’s: we asked eight Czech experts on infectious diseases to rank those types from the most contagious to the least contagious one. Consequently, we evaluated their responses by Saaty method; the results may be seen in the fourth column of Table 8. The individual pairwise comparison matrices were derived from the rankings - the strengths of preference were given by differences in ranks increased by one (resulting in values from 2 to 8), and the inverted values were used to describe the strengths of non-preference to keep the reciprocity. In line with (Dong et al., 2010) and (Forman and Peniwati, 1998), the geometric mean was used to aggregate all the individual judgements into the group one. All the individual matrices were (almost) absolutely consistent (the used rankings cannot violate neither the axiom of transitivity, nor the axiom of multiplicative consistency, see (Alonso and Lamata, 2006)), but it was necessary to check if the final group evaluation was consistent enough with the original individual judgements. In line with (Dong et al., 2010), we used the geometrical cardinal consistency index (*GCCI*) to do this with the results showing that the aggregated group evaluation is sufficiently consistent with the evaluations of all the experts (the authors of (Aguarón and Moreno-Jiménez, 2003) proposed the threshold 0.37 for *GCCI* when *k >* 4, and the values of |*GCCI*| for our experts vary from 0.01 to 0.23).

**Table 8:**
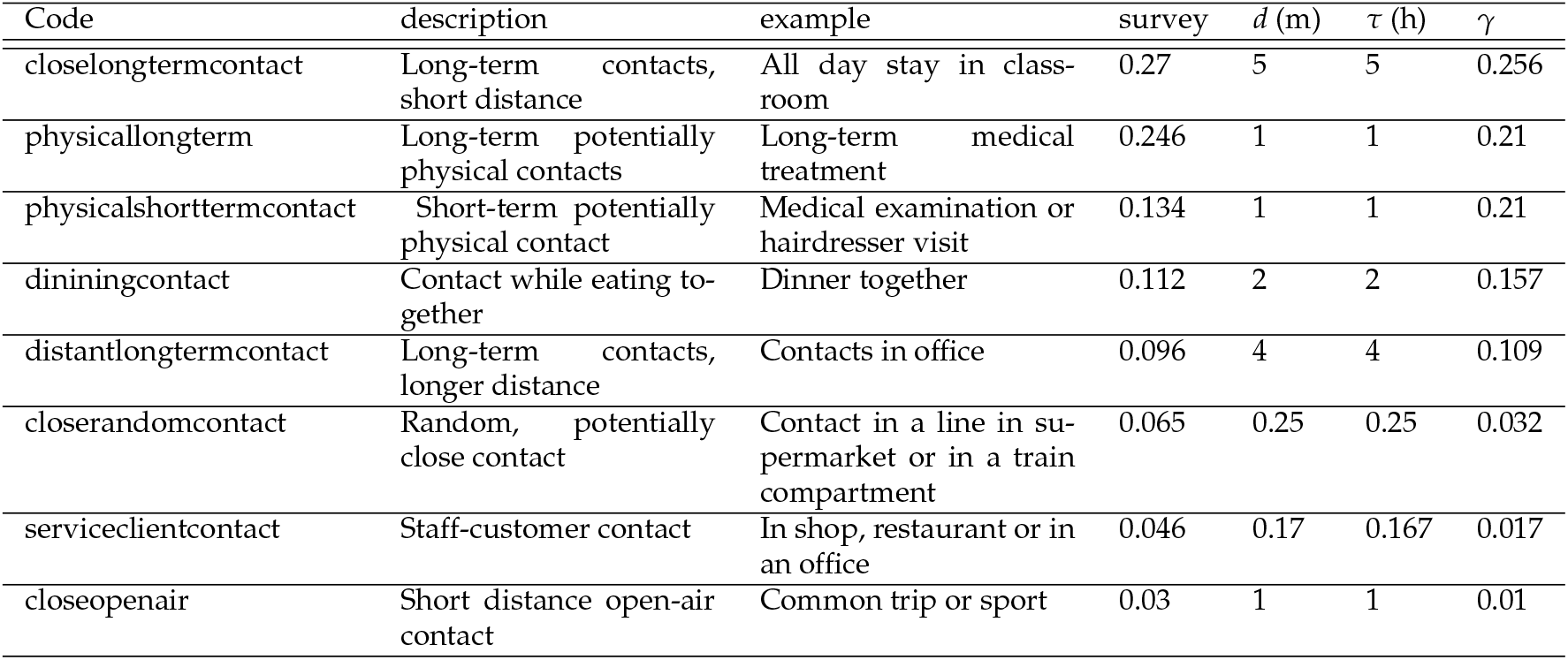
Contact types regarding infectiousness

As the results of our survey can reflect only ordering of the alternatives, not their magnitude, we alternatively evaluate the contagiousness by a simple physical model

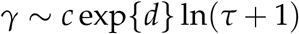

where *d* is a distance of the contact in meters, *τ* is its duration in hours and *c* is a constant which, however, does not matter due to normalization (see above). The values of *d* and *t* were our guesses, reflecting the situations in which we considered the contacts. After minor adjustments of our guesses made for the ordering to agree with the survey, we got final values of *γ*, seen in Table 8.

Which layers were assigned which contagion types can be seen in Table 9.

**Table 9:**
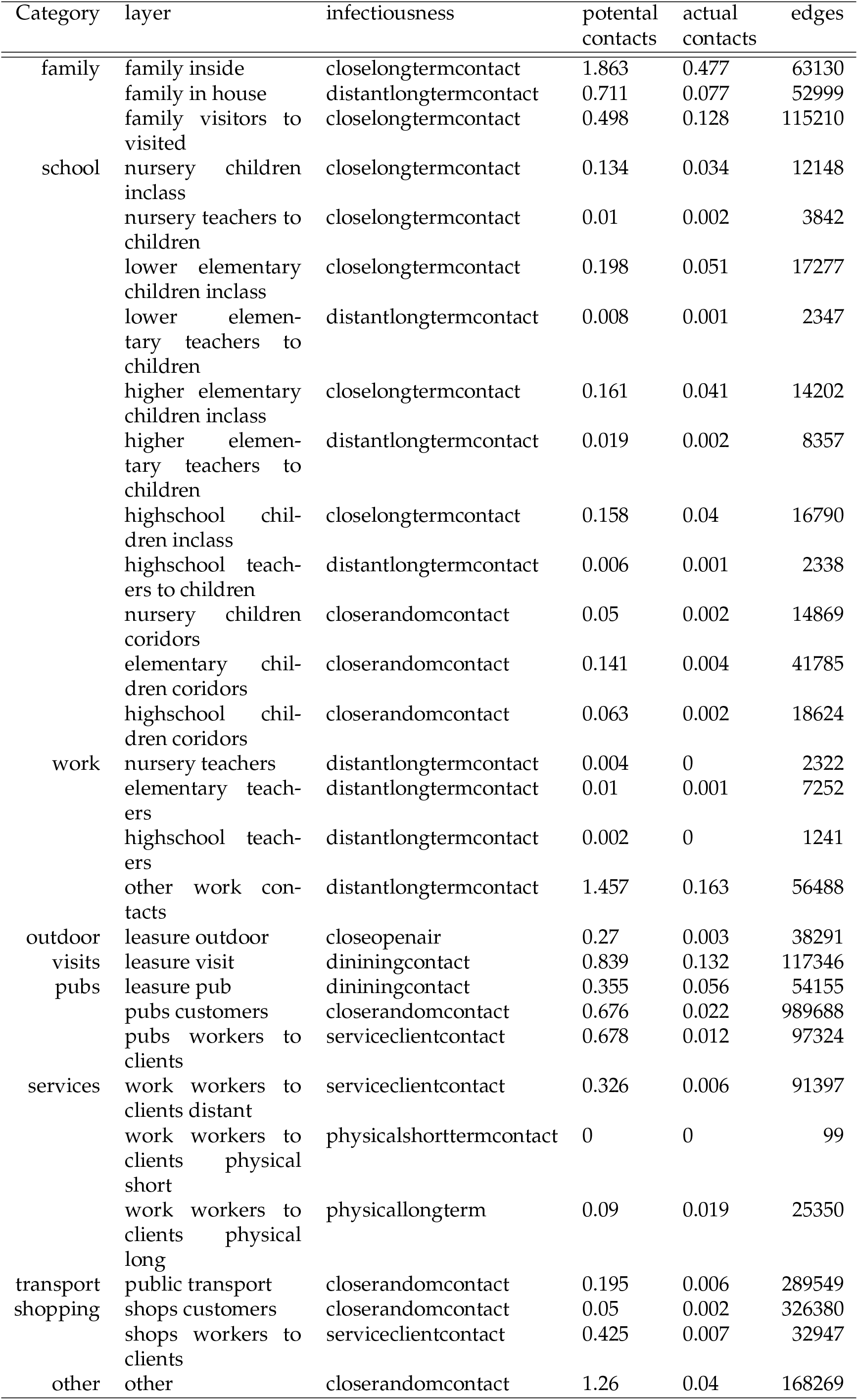
Layers

### 2.6 Simulating interventions

It is no doubt that any working epidemiological model has to take the adaptation of people to the epidemics. Our model enables simulating various interventions via modifications of graph or model parameters. In general, the interventions are of three types – global contact restrictions (site closures, etc.), beta reductions (protective meassures such as masks, proper hand washing) and quarantine/isolation of individuals (including contact tracing).

#### 2.6.1 Contact Restrictions

The data on which we base our contact model, such as the contact matrices from (Prem et al., 2017) or the survey (MEDIAN, 2019), come from the times before the COVID pandemics, when the values of interest clearly differed from the epidemic a probably also post-epidemic times. Moreover, these data changed according to the situation during the epidemic. To incorporate these changes, we used surveys (MEDIAN, 2020) and (PaQ, 2020). The survey (MEDIAN, 2020) compares numbers of contacts of different types between January and the first week of April 2020. The survey (PaQ, 2020), on the other hand, is longitudinal, asking the same set of questions each week of the pandemics. While (MEDIAN, 2020) responds directly to the question what number of contacts of different types people had before the pandemics and during it, it is only one-time survey, speaking only about one time instant in the beginning of April, the study (PaQ, 2020) gives data for each week; however, it surveys only the total number of contacts without discerning their types. Yet additional results of the survey give some information connected to the particular types of contacts, they do not directly provide the numbers of contacts; instead, they give proxy information such as number of pub visits.

To quantify the changes for the purpose of the contact model, we proceeded as follows. To each layer, we assigned an item (answer to a question of a survey) from (MEDIAN, 2020) and a proxy from (PaQ, 2020), see Table 11. Thus, for each particular layer, we had the restriction *m* at the fourth week, estimated by (MEDIAN, 2020), and the series of restrictions *n*_1_, *n*_2_, … at the individual weeks, estimated by (PaQ, 2020). The contact restriction of the layer at week *τ* has been computed as

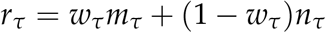

where

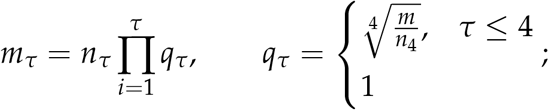

this choice guarantees that, *m*_4_ = *m* and, simultaneously, the information from (PaQ, 2020) is incorporated. Here, *w*_*τ*_ is the weight, such that *w*_1_ = *w*_2_ = *w*_3_ = *w*_4_ = 1 (maximal usage of information from (MEDIAN, 2020)) and *w*_5_ = 0.9, *w*_6_ = 0.8, …, *w*_14_ = 0, *w*_15_ = 0, ….

**Table 10:**
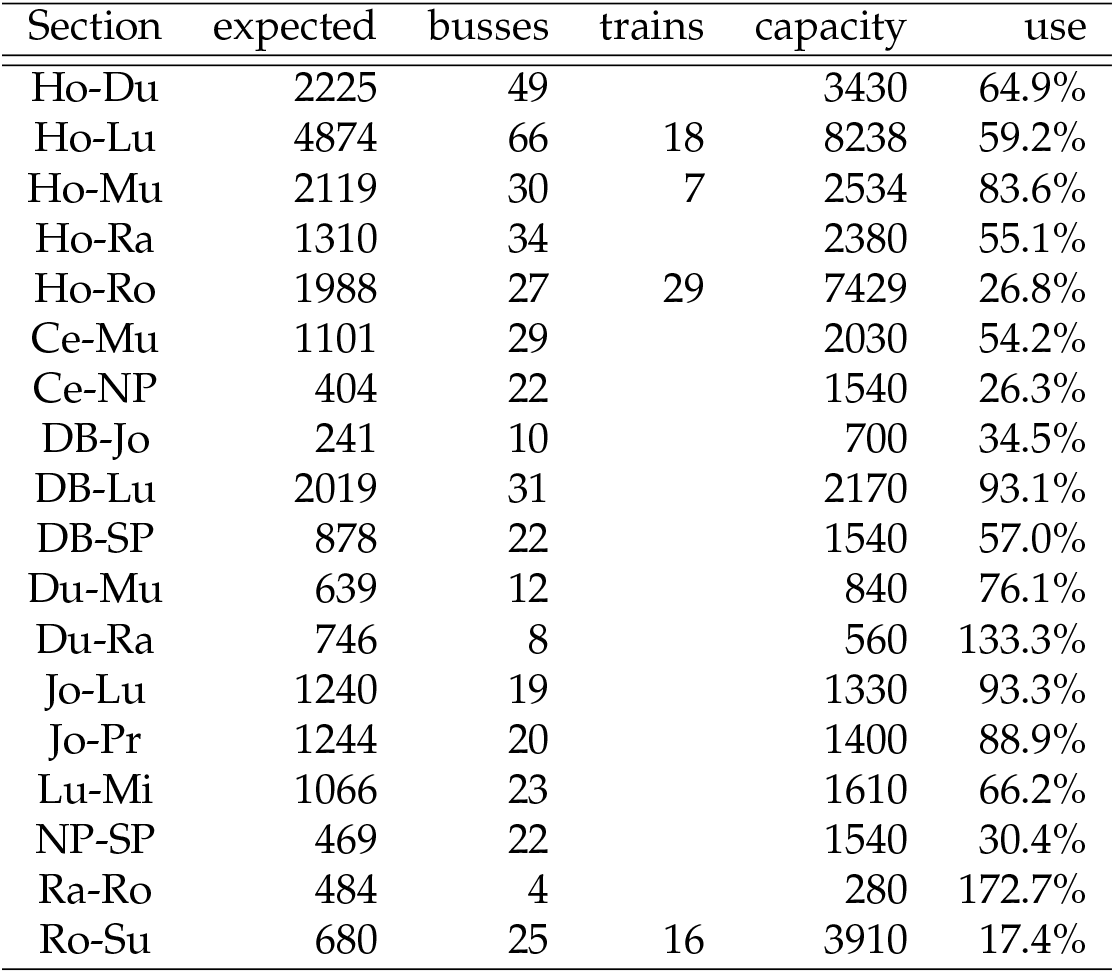
Utilization of public transport.

| Section | expected | busses | trains | capacity | use |
| --- | --- | --- | --- | --- | --- |
| Ho-Du | 2225 | 49 |  | 3430 | 64.9% |
| Ho-Lu | 4874 | 66 | 18 | 8238 | 59.2% |
| Ho-Mu | 2119 | 30 | 7 | 2534 | 83.6% |
| Ho-Ra | 1310 | 34 |  | 2380 | 55.1% |
| Ho-Ro | 1988 | 27 | 29 | 7429 | 26.8% |
| Ce-Mu | 1101 | 29 |  | 2030 | 54.2% |
| Ce-NP | 404 | 22 |  | 1540 | 26.3% |
| DB-Jo | 241 | 10 |  | 700 | 34.5% |
| DB-Lu | 2019 | 31 |  | 2170 | 93.1% |
| DB-SP | 878 | 22 |  | 1540 | 57.0% |
| Du-Mu | 639 | 12 |  | 840 | 76.1% |
| Du-Ra | 746 | 8 |  | 560 | 133.3% |
| Jo-Lu | 1240 | 19 |  | 1330 | 93.3% |
| Jo-Pr | 1244 | 20 |  | 1400 | 88.9% |
| Lu-Mi | 1066 | 23 |  | 1610 | 66.2% |
| NP-SP | 469 | 22 |  | 1540 | 30.4% |
| Ra-Ro | 484 | 4 |  | 280 | 172.7% |
| Ro-Su | 680 | 25 | 16 | 3910 | 17.4% |

**Table 11:**
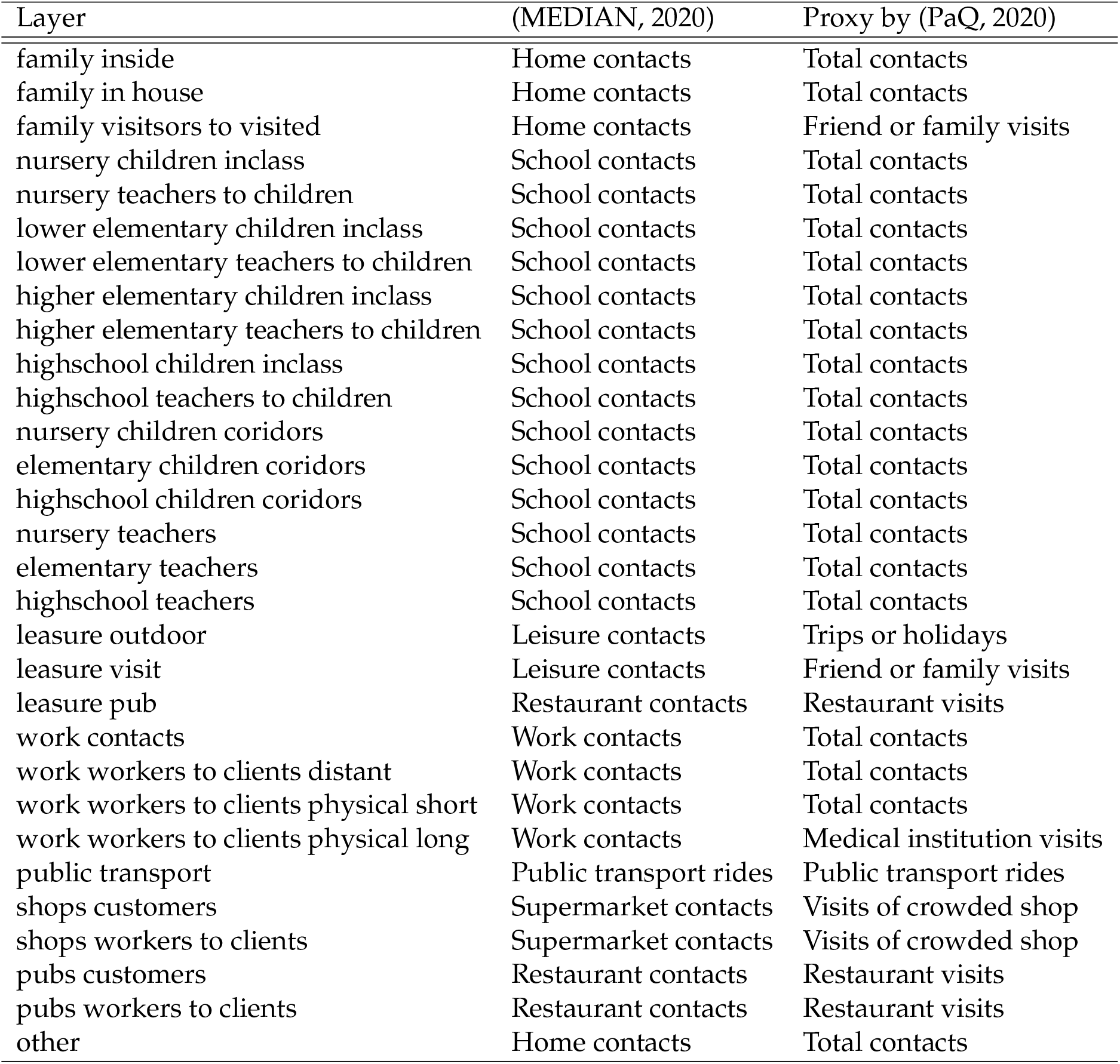
Assigning questions from surveys to layers.

The resulting dynamics of restrictions by category are displayed in Figure 5.

**Figure 5:**
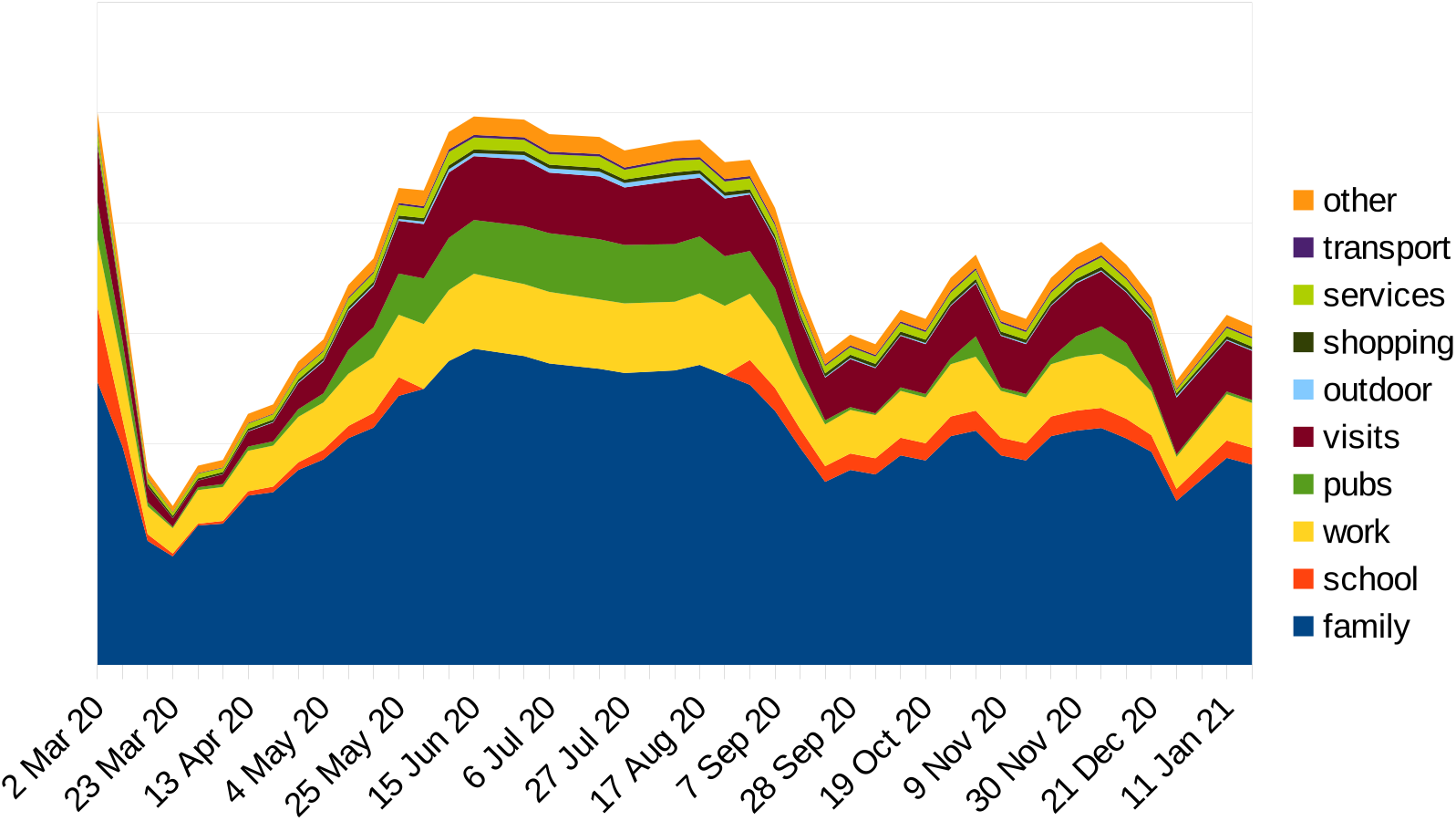
Contact restrictions during the pandemics, contact volumes adjusted for infectiousness.

#### 2.6.2 Beta restrictions

The protective meassures (such as wearing masks, keeping proper distances, frequent hand washing, etc.) adopted by the majority of people during the epidemic reduce the infectiousness represented in model by the parameter *β*. Therefore during simulating the past epidemy flow the parameter *β* is not kept constant but changes in time.

The model uses a calendar of protection levels *ω* (real number between ⟨0, 1 ⟩ representing how much the hygienic protective measures were active). This calendar is crated based on data provided by (PaQ, 2020).

The parameter *β* is modified using the following formula

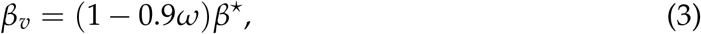

where *β*^*\**^ is a value of a parameter beta while no restrictions are active. This is valid for all edges except edges on layer 1 (families in one household), where the strength of reduction is 20%:

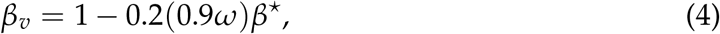

where *v* is an edge on layer 1.

#### 2.6.3 Quarantine and contact tracing

The last type of interventions works on individual nodes. It enables the simulation of isolation or quarantine of individuals and contact tracing. Again, the simulation is based on graph modification, in this case local (the edges of given node that should be isolated are weakened for required period of time).

The policy module responsible for qurantine (called Quarntine Policy (QP)) monitors indiviual node’s states and puts some of nodes to isolation.

Two main types of QP were implemented: self-isolation policy (SIP) and quarantine policy with contact tracing (CTP).

SIP is a simple policy, it simulates the situation when people after developing symptoms stay home (since they are responsible or feel too badly to continue their normal lifestyle). Once a node develops symptoms, it is isolated with a certain probability. It means, that the node is stored in a deposit object for a given number of days (typically 7). For this period, nodes adjacent edges are reduced, i.e. their probabilities are set to zero or weakened (except family edges on layer 1 – family). After the duration time in deposit object passes and the node leaves the state *I*_*s*_, the original probabilities of edges are recovered (unless they are reduced by other quarantine at the same time).

Another situation is that a node is detected and is forced to stay in isolation. This is covered by CTP that tracks detected nodes, puts them to isolation, and simulates a contact tracing.

CTP is described in Alg. 1 and its basic workflow sketched at Fig. 6. The main purpose of this algorithm is to track detected nodes and their contacts and keep them in isolation or quarantine for a given number of days (in CR during spring 14, later changed to 10). As was valid during spring in the Czech Republic, two negative tests are needed to end up the isolation/quarantine. Isolation and quarantine are again modeled by reducing probabilities of edges adjacent to quarantined or isolated nodes.

**Figure 6:**
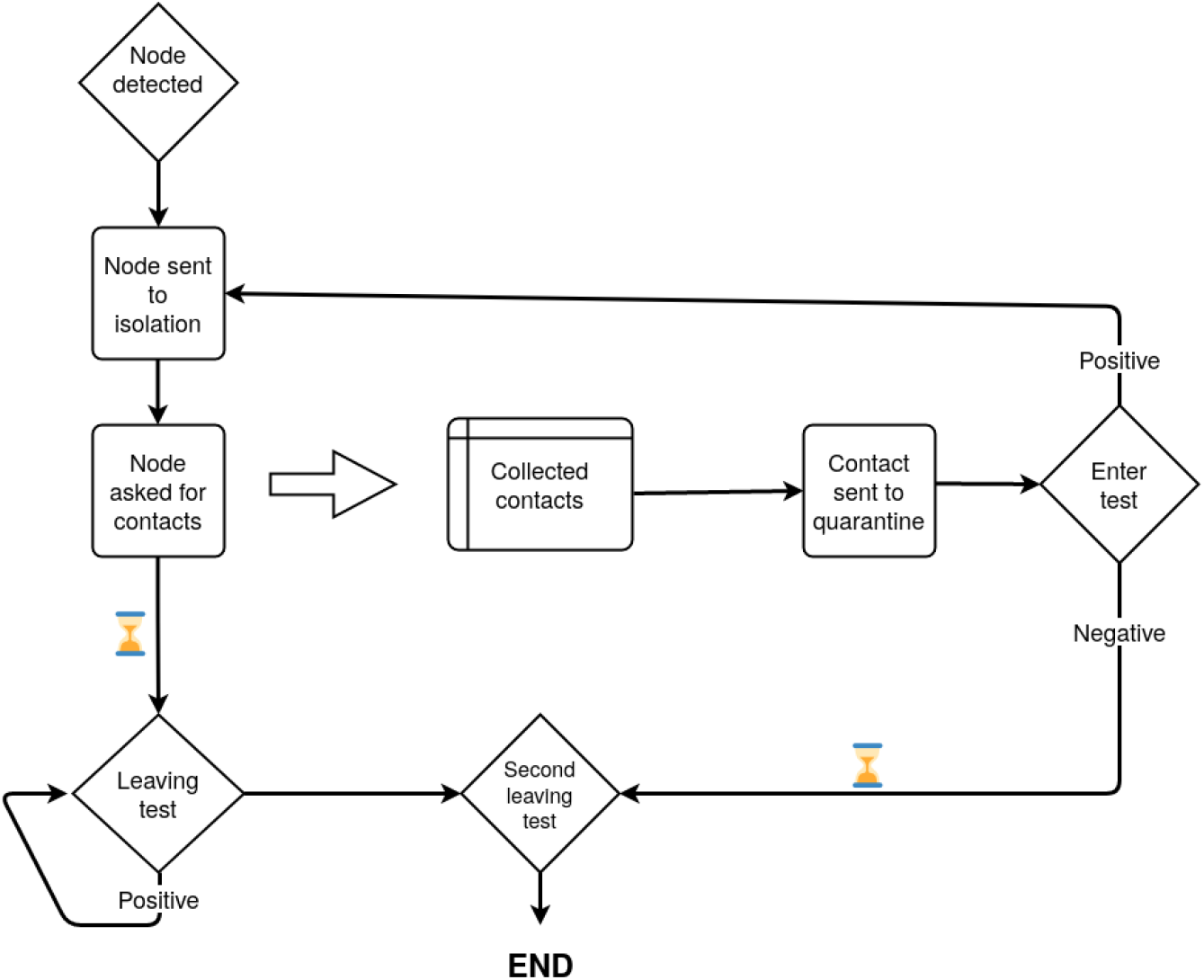
Quarantine and contact tracing workflow.

Once a node is detected, it is put in isolation and is contacted and asked for contacts (there is a delay when the node waits for the phone call). These contacts are put to quarantine and have to undergo an enter test (usually in 3 to 5 days). If the enter test is negative, it has to pass another negative one at the end of its quarantine period. If the result is positive, the node is put in isolation and is also contacted and asked for contacts.

The contact is a node adjacent to an interviewed node on the condition that the connecting edge was active (see Sec. 2.2) in a given number (typically 5) of days and with the probability equal to the riskiness of the corresponding edge’s layer type.

To simplify and formalize this concept, the CTP is defined by a tuple (*p*1, *p*2, *p*3, *p*4). The *p*_*i*_ corresponds to layer type (see Table 12) and defines the probability that a contact on given layer will be collected (i.e. successfully traced). For example, if contact tracing (1, 0, 0, 0) is used, all contacts connected by family edges are collected (recalled), others are ignored. For riskiness (1, 0.5, 0, 0) all family contacts are collected, school and work contacts are recalled with probability 0.5 and the rest of contacts is ignored.

**Table 12:** Four groups of contacts with different recall rates based on tracing policies.

| Notation | Layer |
| --- | --- |
| p1 | family (layers 1-3) |
| p2 | school and work (layers 4-17 + 21-24) |
| p3 | leisure (layers 18-20) |
| p4 | other (layers 25-30) |

## 3 Results

The software implementation of the previously described model M is realized in Python, and it is publicly available at https://github.com/epicity-cz/model-m/releases. Figure 7 presents a scheme of software modules of the model.

**Figure 7:**
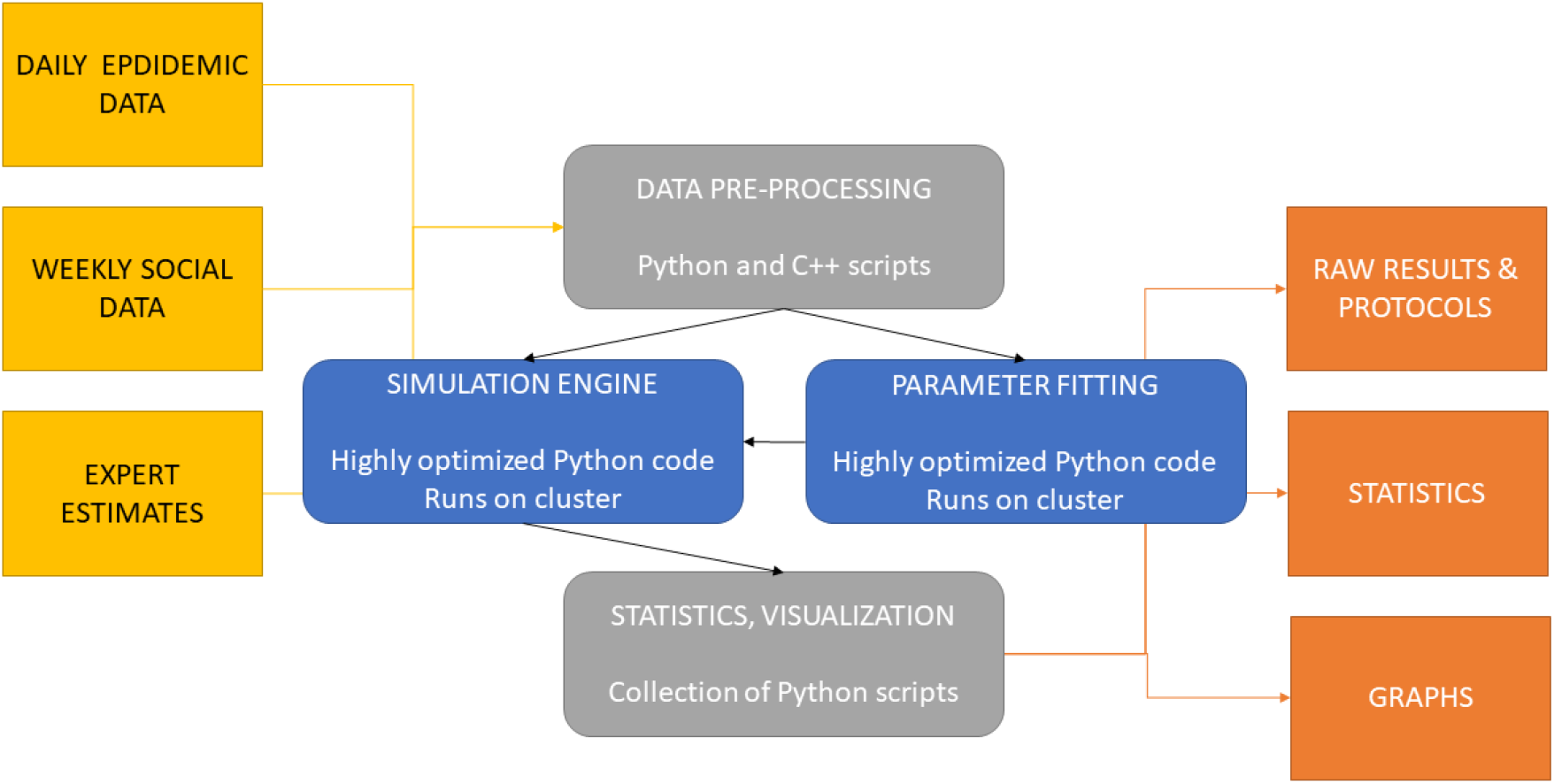
Scheme of a software implementation of model M.

As an illustration of the model usage we present an example where the parameters of the model were fitted to the situation in Czech Republic in Spring and Summer of 2020. Typical output of the simulation contain the numbers of active and detected cases per day, together with further context dependent outputs, such as numbers of tests, etc. Figure 8 illustrates a result of such an experiment. Figure 9 presents a geographic visualization of one simulation run.

**Figure 8:**
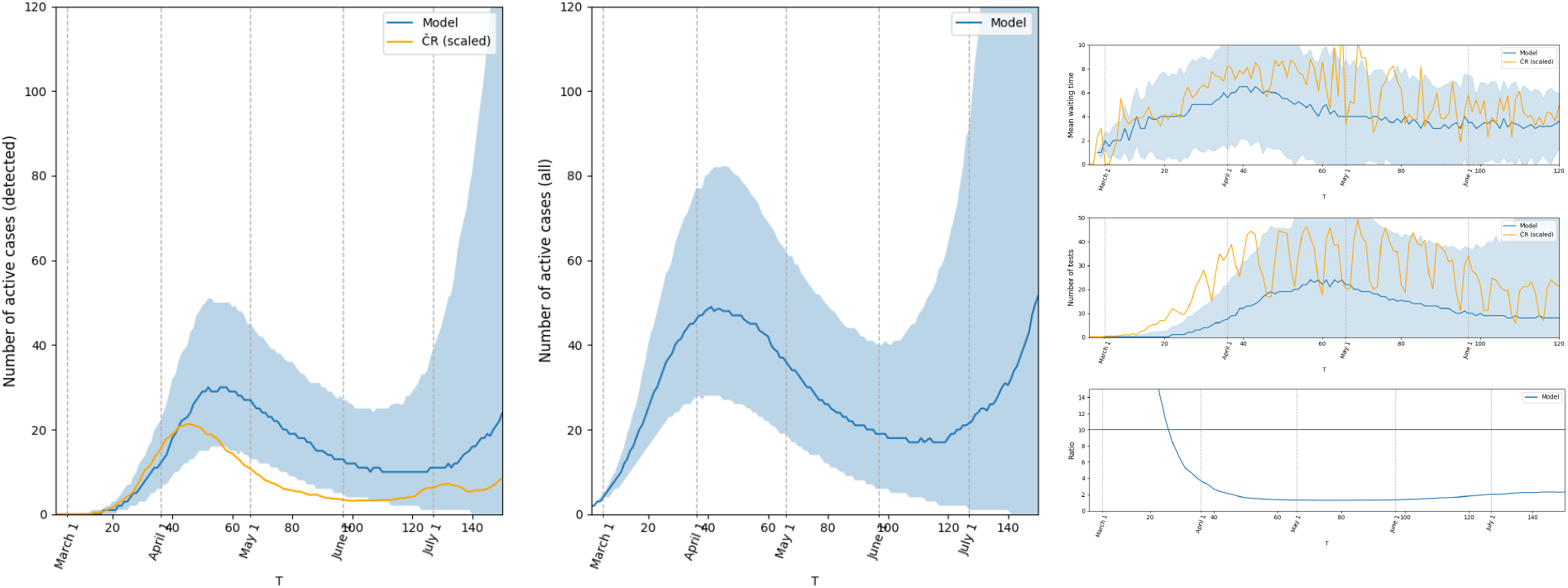
The fit of our model to the situation in the Czech Republic. Left: detected active cases (sum of numbers of nodes in detected states, i.e. *E*_*d*_, *I*_*dn*_, *I*_*da*_, *I*_*ds*_, *J*_*dn*_, *J*_*ds*_). Center: all active cases (sum of all *E, I*_*a*_, *I*_*s*_, *I*_*n*_, *J*_*n*_, *J*_*s*_ plus their detected counterparts). Right top: Average times between the first symptoms and the test result. Right middle: Number of all tests (should be always underestimated in a model, we do not realise all tests, since we do not care about negative ones except those in quarantine)). Right bottom: Ratio of all active cases and detected active cases. Number 1.0 stands for all detected, *>* 1.0 undetected ill nodes.

**Figure 9:**
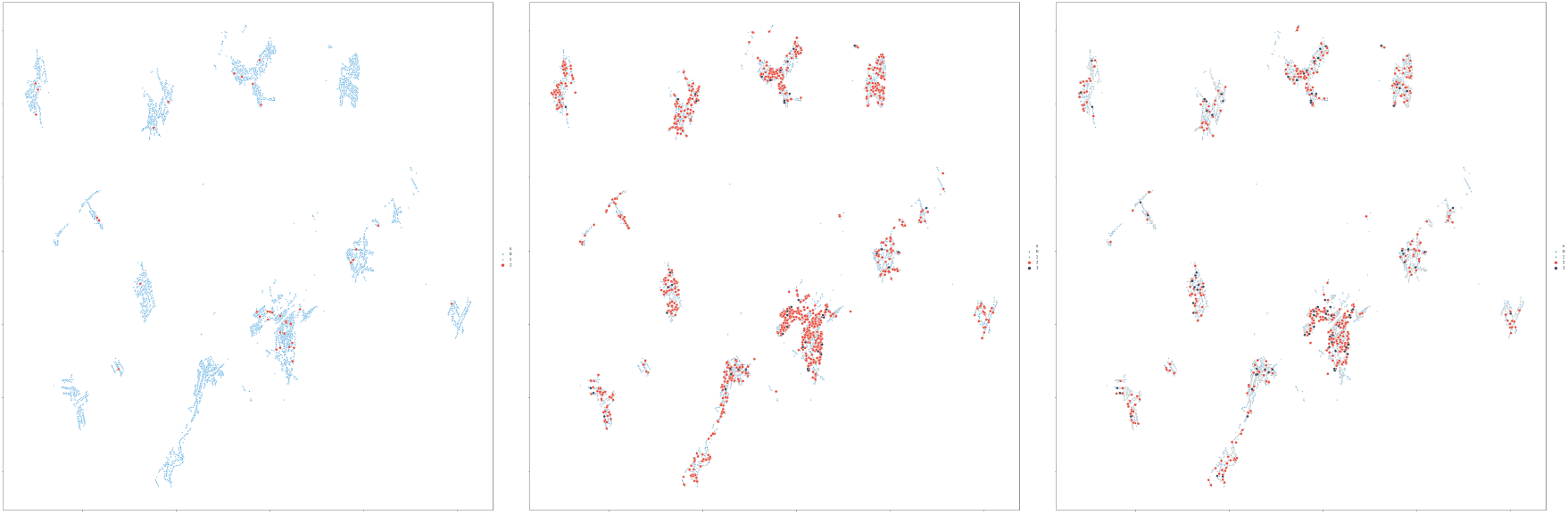
Geographic visualization of one simulation run.

### Algorithm 1 Main Routine – Contact Tracing Policy (CTP). See also Alg. 2 for the description of auxiliary routines

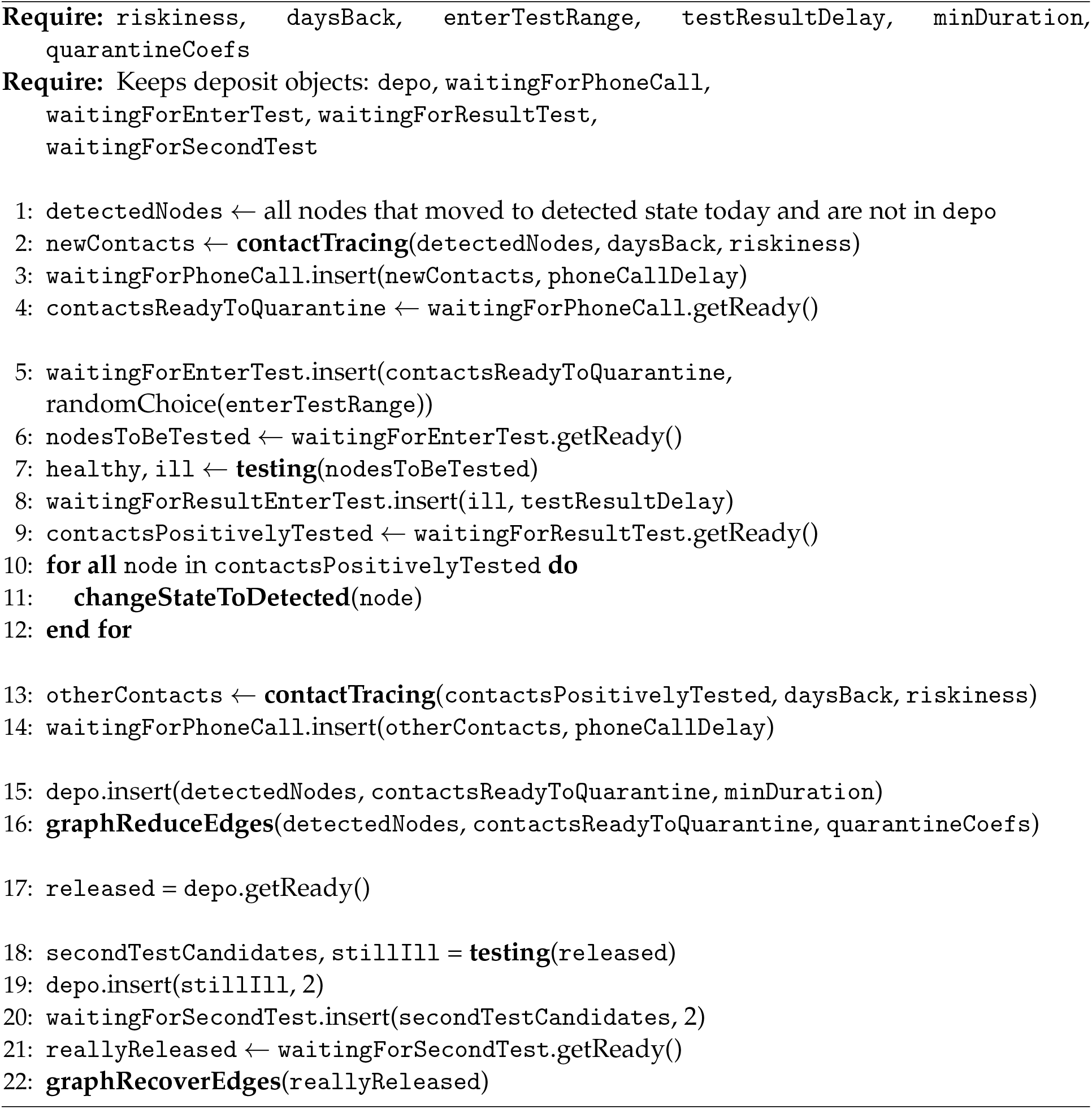

### Algorithm 2 Auxiliary Routines – objects and functions used in Alg. 1

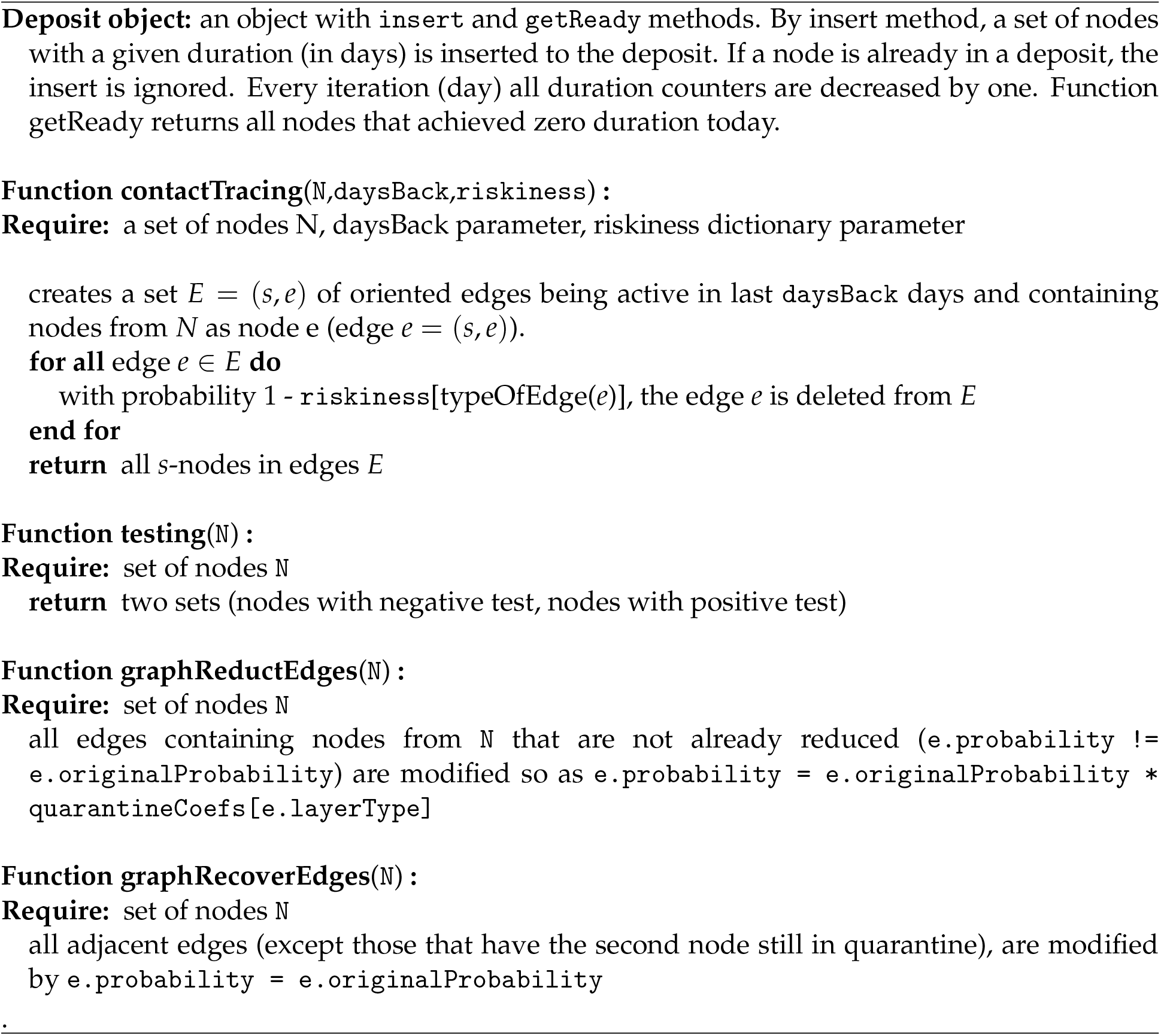

## 4 Discussion

We have presented a technical description of an agent-based model of the Hodonín municipality of the Czech Republic. The model is unique in its detail in which the synthetic population corresponds to the real population of the area.

The second important feature is a detailed graph of social network of contacts. The contacts are divided into 30 layers corresponding to families, households, workplaces, schools, shops, restaurants, and public transportation. They are modeled as closely as possible using various data resources in order to obtain a faithful contact network.

Moreover, we have a detailed calendar of changes of the behavior of Czech Republic inhabitants from an ongoing sociological survey. This allows to dynamically change the contact rates depending on real situation and to model the effect of various interventions.

The interventions in our model include non-pharmaceutical individual measures, such as personal protection and distancing, testing, isolation and quarantine procedures with limited resources, and tracing policies with varying efficiency in different contact types.

The software implementation is rather general, given the right graph, it can be easily used for different regions, but also to environments such as schools or workplaces, in order to compare the efficiency of specific epidemics measures.

Many software packages for epidemiological simulations have emerged in previous year, such as the extensive suite Covasim (Kerr et al., 2021) that allows to run simulations on random networks as well as synthetic populations. Our solution is similar in the modeling approach and the range of implemented interventions. What makes our Model M unique is the level of detail in the population and contact network structure.

## Data Availability

Data and code is available in the link provided.

https://www.github.com/epicity-cz/model-m/releases

## Footnotes

* To see it, note that the mean number of contact is computed as the mean of the sum of indicators of the individual contacts.

† When the number of potential attendees is large, namely larger than 25, we, instead linking all of them, created a limited number *k* of contacts for each individual, scaling the probabilities accordingly. Different constants *k* are chosen in different situations.

‡ If there is (are) another household(s) in the apartment of the parents, we assume it (them) be the children’s one(s) and we sought for accordingly less number of external households.

§ The distance was computed as a sum of square differences of the the matrix elements, weighted by population fractions corresponding to the column and the row. After finding optimal *p*’s, these values were scaled in order the expected contact numbers implied by both matrices to match.

¶ The probability of the coin, obtained from the team running (CDV, 2020), was 0.65/0.18/0.19 for a school/work/other trip.

∥ The capacities were computed by means of online timetables and a fan server vagon.cz.

